# Chromatin, transcriptional and immune dysregulation in children with neurodevelopmental regression

**DOI:** 10.1101/2025.03.05.25322433

**Authors:** Hiroya Nishida, Velda X Han, Brooke A Keating, Katherine G Zyner, Brian Gloss, Nader Aryamanesh, Pinki Dey, Lee L Marshall, Erica Tsang, Sarah Alshammery, Xianzhong Lau, Ruwani Dissanayake, Shekeeb Mohammad, Mark Graham, Shrujna Patel, Russell C Dale

**Author notes:** Corresponding author: Prof Russell Dale Kids Neuroscience Centre, The Children’s Hospital at Westmead, Faculty of Medicine and Health, University of Sydney, Sydney, New South Wales, Australia The Children’s Hospital at Westmead Clinical School, Faculty of Medicine and Health, University of Sydney, Sydney, New South Wales, Australia, The Brain and Mind Centre, The University of Sydney, Sydney, New South Wales, Australia. Joint first author (Hiroya Nishida, Velda X Han). Joint senior author (Shrujna Patel, Russell C Dale).

## Abstract

Neurodevelopmental including autistic regression in children is often associated with inconclusive diagnostic investigations, leaving the underlying pathophysiology largely unexplored and poorly understood. To address this, we conducted peripheral blood multi-omics profiling in 15 children with neurodevelopmental regression (median age 9 (4-14) years, 60% males), and 15 healthy controls (median age 12 (7-16) years, 60% males). Our comprehensive analysis included bulk RNA sequencing (8 regression vs. 8 controls), single-cell RNA sequencing (two cohorts of 4 regression vs. 4 controls), bulk proteomics (4 regression vs. 4 controls), and phosphoproteomics (4 regression vs. 4 controls). Despite clinical heterogeneity, we identified convergent pathophysiological processes, including dysregulated pathways in epigenetic pathways (chromatin remodelling, histone modifications, transcription factors) and immune responses, as well as downregulated ribosomal and translational pathways. These disruptions in epigenetic, immune and translational processes may arise from complex genetic and environmental interactions, offering opportunities for therapeutic interventions that specifically target these pathways.

## Introduction

Neurodevelopmental disorders (NDDs) affect 1 in 6 children, with rising prevelance^1^. These disorders arise from disruptions in brain development during critical prenatal and postnatal developmental windows, influenced by both genetic and environmental factors^2^. Key developmental processes, including neurogenesis, synaptogenesis, myelination, and synaptic pruning by microglia, actively shape brain architecture during this period^3^. While genetic factors play a critical role in these processes, the absence of a single pathogenic gene variant in many affected individuals highlights the contribution of common gene variants^4^. Genes involved in chromatin modification, transcriptional control, synaptic organisation, and immune regulation have been implicated in the pathogenesis of NDDs^5–7^. Additionally, environmental triggers also significantly impact these developmental processes, leading to changes in brain structure, cognitive functions, and behaviour^2^. The interplay between genetic and environmental factors is complex and proposed to operate through epigenetic modifications and immune pathways^2^.

Developmental trajectories in children with NDDs are classically characterized by gradual and steady progress. However, some children experience regression, defined by the loss of previously acquired skills. This regression can be abrupt, triggered by specific events, or gradual in nature. Autistic regression, observed in up to one third of individuals with autistic spectrum disorders (ASD), typically manifests in the second year of life^8–12^, although sometimes occurs at an older age, referred to as “childhood disintegrative disorder”. Autistic regression involves losing previously acquired skills in social interaction, language, cognition, or behavioural changes^8–12^. In addition, some children lose developmental skills, such as cognitive decline, memory loss or executive dysfunction, without fulfilling the criteria for autism or a definable childhood dementia syndrome. Despite being widely recognised in the literature, the pathophysiology of autistic and neurodevelopmental regression remains unclear in a significant proportion of patients. For example, the current diagnostic yield of exome sequencing in cases of autistic regression is only 9%^13^. Likewise, neuroimaging and cerebrospinal fluid testing is typically normal or unhelpful. It is therefore believed that most cases are not attributable to a rare de novo or inherited DNA variation, but rather a combination of common vulnerability genes and environmental factors^14^. Unfortunately, treatment in neurodevelopmental regression remains limited, primarily focussed on managing comorbid behaviours with anti-psychotics, stimulants and anti-depressants, and providing developmental support with speech, occupational, physical and psychological therapies.

We hypothesize that epigenetic and immune dysregulation in both the blood and brain, occurring during critical developmental periods, plays key roles in neurodevelopmental regression. In this study, we investigated children with neurodevelopmental regression with unexplained aetiology using bulk RNA sequencing (RNAseq), single-cell RNA sequencing (scRNAseq), proteomics, and phosphoproteomics of peripheral immune cells and found evidence of chromatin, transcriptional and immune dysregulation. These insights provide opportunities for biomarkers and future therapeutics targeting epigenetic and immune pathways.

## Methods

### Participant selection

Children (<18 years of age) were recruited into this study if they met criteria for a prior history of neurodevelopmental (autistic or cognitive) regression.

1. Regression was defined as one or more episodes of losing previously established skills; the loss of spoken language (such as a child who was regularly speaking phrases who stops speaking) and/or loss of social and communication abilities (such as a child previously using gestures and making regular eye contact who loses such social responsiveness^15^), or cognitive decline/memory loss. In all included children, there was a clear history of loss of previously acquired neurodevelopmental or cognitive skills, associated with functional decline- corroboration of the parental observations from teachers and other family members was sought in all patients.
2. Normal investigation using routine application of neuroimaging, cerebrospinal fluid and genomic testing (as defined by Australian Medicare supported genomic testing). The investigation was determined by the treating clinicians.
3. All children had ongoing symptoms and were in the ‘chronic phase’ of their illness. No patient had an infectious disease in the preceding two weeks prior to blood sampling.
4. The patients who had scRNAseq and proteomics/phosphoproteomics (n=8) were all screened for rare pathogenic variants that cause autistic spectrum disorder (trio exome n=3, and autism gene panel n=5), and were negative.

### Control selection

The inclusion criterion for healthy controls was absence of neurodevelopmental disorders, autoimmune diseases, severe allergic conditions, as well as absence of infection two weeks before blood draw. Controls were sex- matched to patients, and there was no significant age difference between patients and controls.

### Semi-structured clinical interview

Clinicians recorded the child’s demographics, family history, birth history, comorbidities and medication history after a semi-structured clinical review for all participants, using DSM-5 criteria^16^. Details around the regression episodes were collated including development prior to regression, timings of regression, and developmental domains affected, including loss of social communication, loss of language/meaningful words, loss of imaginative play/restricted play, and restricted/repetitive patterns of behavioural interest. Videos were used to corroborate parental observations of loss of developmental skills.

### Sample collection

After written consent, venous blood samples were collected for testing in PAXgene™ blood RNA tubes (Qiagen, Hilden, Germany) and Acid Citric Dextrose tubes (BD). Peripheral blood mononuclear cells (PBMCs) were isolated from the whole blood within 12 hours of sample collection using a Ficoll density medium (Cytiva) in SepMate™ tubes (Stemcell Technologies). PBMCs were cryopreserved in 10% dimethyl sulfoxide (Amrescro, WN182-10ML) in foetal bovine serum (FBS, Thermo Fisher Scientific) and stored at -80°C in controlled cooling chambers for 24 hours before being transferred to liquid nitrogen for storage until single cell RNA-sequencing.

### Bulk RNA sequencing

Bulk RNA sequencing was performed in 8 children with neurodevelopmental regression compared to 8 healthy controls (Figure 1, refer to Table S1). This workflow includes RNA extraction from PAXgene™ blood RNA tubes, depletion of ribosomal RNA via hybrid capture (Illumina Ribo-Zero), and Illumina TruSeq Stranded Total RNA Library Preparation (input 200-1,000 ng of Total RNA). The stranded RNA samples were sequenced on the Illumina NovaSeq 6000 next generation sequencing platform (2 x 150 base pairs) for a depth of 50 million paired end reads. The cleaned sequence reads were aligned against the *Homo sapiens* genome (Build version hg38), and the STAR aligner (v2.5.3a) was used to map unique reads to the genomic sequences^17^.

**Figure 1.**
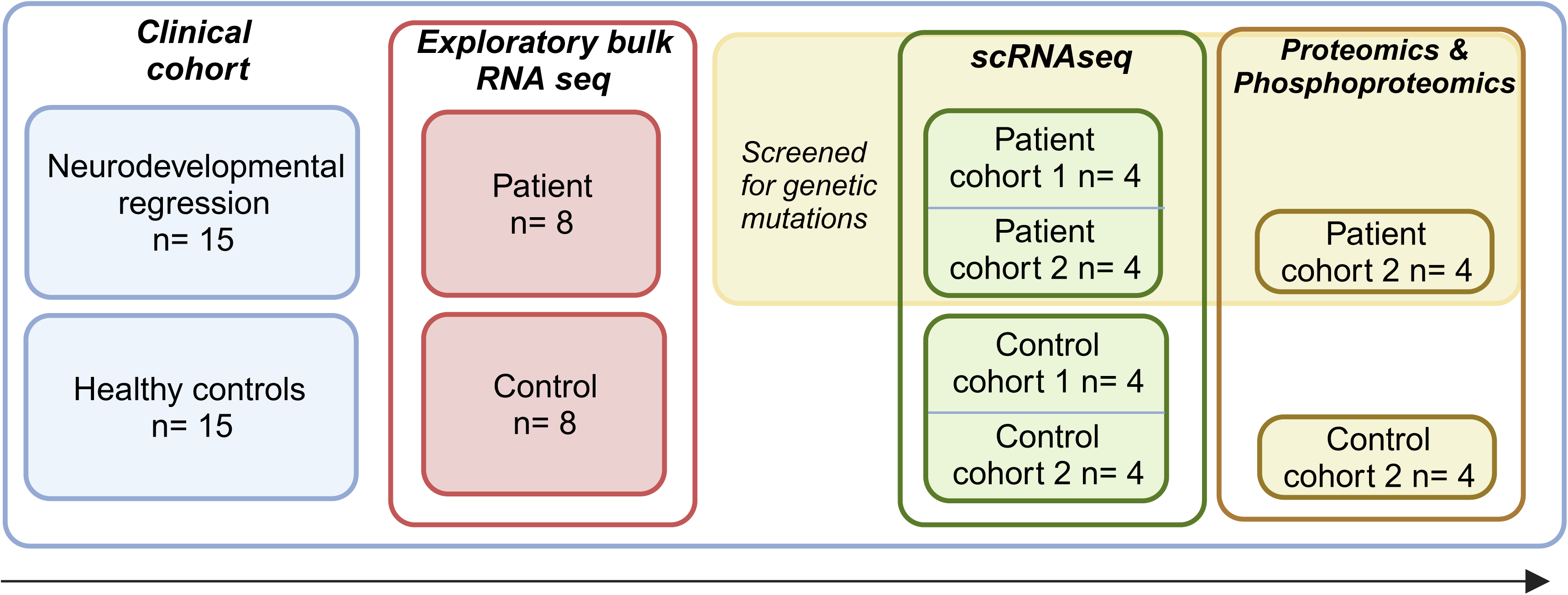
**Study workflow** 15 children with neurodevelopmental regression were recruited for this study and underwent comprehensive clinical assessments. Exploratory bulk RNA sequencing was performed, comparing samples from 8 affected children to 8 healthy controls. Genetic screening of 8 of the children with neurodevelopmental regression who were used in scRNA sequencing and proteomic/phosphoproteomic studies (1 overlap with the initial group) revealed no pathogenic mutations. These 8 children were further analyzed using single-cell RNA sequencing in two separate cohorts, each comprising four affected children and four matched controls. Additionally, the second cohort (n=4) underwent proteomic and phosphoproteomic analyses to further investigate the underlying molecular mechanisms.

### HIVE™ Single-cell RNA sequencing

Single-cell RNA sequencing was performed in 8 children with neurodevelopmental regression compared to 8 healthy controls (2 cohorts of 4 patients vs 4 healthy controls). Red blood cells (RBC) were depleted from whole blood by immunomagnetic negative selection, using the EasySep™ RBC depletion reagent protocol (catalog #18000), preserving all leukocyte populations, including granulocytes. Two cohorts of 8 HIVE devices (4 patients and 4 controls for each cohort) were each loaded with 30,000 cells in 1 mL of DPBS + 1% FBS, followed by 3mL of cell media (DPBS + 1% FBS), according to the manufacturer’s protocol. The leukocytes were loaded into the HIVE collector within an hour of blood sampling from all individuals to minimise neutrophil activation. Single-cells settled into picowells of the HIVE containing 3’ transcript-capture beads. HIVEs were then placed onto a spin plate and spun at 30 x G for 3 minutes at room temperature. Media was removed from the HIVE, and 2 mL sample wash solution was added to each, following the manufacturer’s instructions for the HIVE sample capture kit (Honeycomb Biotechnologies). The sample wash solution was removed, and the cell-loaded HIVEs were frozen at -80°C after adding 1 mL of the cell preservation solution, before being transferred to the Australian Genome Research Facility (AGRF Ltd) and processed through to single-cell NGS libraries.

HIVE devices were processed according to the manufacturer’s instructions. Briefly, cell-loaded HIVE devices were sealed with a semi-permeable membrane, enabling the use of the strong lysis solution and the addition of a hybridization solution. Beads with captured transcripts were extracted from the HIVE device by centrifugation.

Subsequent HIVE library preparation steps were performed in a 96-well plate format. The size distribution of the final amplified libraries was determined using the TapeStation and the final concentrations of the pooled libraries were accurately quantified using the qPCR assay. HIVE scRNAseq libraries were paired-end sequenced (2 × 150 bp) with custom primers (provided in the HIVE single-cell RNA seq processing kit) on an Illumina NovaSeq 6000 platform.

### Proteome analysis and preparation from PBMCs

Proteomics and phosphoproteomics were performed in 4 children with neurodevelopmental regression and 4 controls (second cohort who had scRNAseq). Following isolation, PBMCs were lysed in 200 µL lysis buffer containing 0.8% Triton X-100, PhosSTOP, 50 mM HEPES (pH 7.4, adjusted with NaOH), and 2 mM phenylmethylsulfonyl fluoride (PMSF, in ethanol). The lysates were heated at 85°C for 10 minutes in a heating block and stored at -80°C. The samples were thawed, benzonase treated, reduced, alkylated, precipitated and digested as described previously^18^. Approximately 90 µg of peptide from each sample was labelled with a TMTpro reagent and combined. A portion of the combined sample was fractionated by hydrophilic interaction liquid chromatography on a Vanquish Neo HPLC system (Thermo Fisher Scientific) with a 250 mm long and 1 mm inside diameter TSKgel Amide-80 column (Tosoh Biosciences). The HILIC gradient used 90% acetonitrile, 0.1% TFA (Buffer A) and a solution of 0.1% TFA (Buffer B). The flow rate was 50 μL/min. The gradient was from 100% Buffer A to 60% Buffer A in 35 min. Fractions were collected into a 96-well plate using an FC204 fraction collector (Gilson) at 1 min intervals, monitored by absorbance of UV at 214 nm. UV signal was used to combine select fractions into similar amounts of peptide. Fractions were dried and reconstituted in 0.1% formic acid. The LC-MS/MS was performed as described previously^18^ using a Dionex UltiMate 3000 RSLC nano system and Q Exactive Plus quadrupole-orbitrap mass spectrometer (Thermo Fisher Scientific). We obtained 5,961 proteins suitable for analysis.

### Phosphoproteomics analysis

Phosphopeptides were enriched using a previously described method^19^. Hydrophilic interaction liquid chromatography was used to fractionate the phosphopeptides as for the proteomics. LC-MS/MS was performed as for the proteomics except that gradient was from 1% buffer B to 5% buffer B in 1 min, to 25% buffer B in 74 min, to 35% buffer B in 8 min, to 99% buffer B in 1 min, held at 99% buffer B for 2 min, to 99% buffer A in 1 min and held for 8 min as the flow rate increased to 275 nL/min.

The MS/ settings were the same as for proteomics except that maximum ion time was 115 ms and the normalized collision energy was 34. We obtained 7,550 phosphopeptides suitable for analysis.

### Proteomics and phosphoproteomics database searching

The raw LC-MS/MS data was processed with MaxQuant v1.6.7.0 using the following settings: variable modifications were oxidation (M), acetyl (protein N-terminus), deamidation (NQ) and phospho (STY); carbamidomethyl (C) was a fixed modification; digestion was set to trypsin/P with a maximum of 3 missed cleavages; the TMTpro correction factors were entered for lot XA340093; minimum reporter peptide ion faction was 0.6; the *Homo sapiens* reference proteome with canonical and isoform sequences downloaded Dec 6 2023 with 80,685 entries and 20,596 genes; the inbuilt contaminants fasta file was also used; minimum peptide length was 6 and maximum peptide mass was 6,000 Da; second peptides search and dependent peptides search were enabled; all modified peptides and counterpart non- modified peptides were excluded from protein quantification. All other settings were default.

### Bioinformatic analysis Analysis of omics datasets

Omics data were analyzed in the R statistical environment^20^ with tidyverse. For bulk RNA sequencing and proteomics, filtering and normalization steps were first performed. Subsequently, normalization with removal of unwanted variation, via the remove unwanted variation *(RUV)* R package was performed ^21^. RUV is commonly used in human samples to better discern biologically relevant variations associated with the disease of interest, particularly in the presence of significant inter-sample biological diversity^22^. This method relies on having a set of endogenous negative control genes known *a priori* not to be differentially expressed with respect to the biological factor of interest ^21,22^. For this study, a set of 500 empirical negative control proteins with little or no change in RNA expression across samples was identified from an initial ANOVA test. In the bulk RNA sequencing (k=10), proteomics (k=4), and phosphoproteomics (k=4) (factors of unwanted variation) were used respectively to remove genes that had minimal differential expression, compared to negative control genes. For linear modelling, the limma R package was used and the p-values were calculated using the empirical Bayes method ‘eBayes’ function^23^. The false discovery rate correction was applied to the p-values by calculating the adjusted p-values. Significant differentially expressed genes/proteins were defined as those with adjusted p-values/false discovery rate (FDR) < 0.05.

For single cell transcriptomics, the data was analysed using the Seurat package^24^. Cells with a high mitochondrial transcript ratio (>0.15) were excluded. Experiments were integrated using the *FindIntegrationAnchors* function in Seurat then immune cell types were assigned using *scPred*^25^. Merged data were then split by cell type and separately normalized, scaled, integrated between patient using harmony^26^, then UMAP (uniform manifold approximation and *projection)* projections were made using the first 30 dimensions. Differentially expressed genes with significant FDR values <0.05 were identified using *FindMarkers*.

#### Pathway enrichment analysis

*Gene set enrichment analysis (GSEA) for bulk RNAseq, scRNAseq and proteomics*: The genes and proteins were ranked based on their sign(logFC) x log10Pvalue scores ^27,28^. Enriched gene sets were identified based on a running sum statistic (normalized enrichment score (NES)) and statistical significance, based on the false discovery rate (FDR). Significant GSEA GO pathways (FDR <0.05) were further simplified using the *simplify* function in clusterProfiler^29^.

*Over representation enrichment (ORA) analysis for phosphoproteomics:* We took an approach that deliberately reduced the significant pathways identified by (a) limiting the background to only proteins we could detect by mass spectrometry and (b) using the ranked list method within gProfiler, which weights enrichment toward highly ranked genes. We separated up- and down-regulated phosphorylation, assigned the maximum positive and negative quantitative values for phospho-regulation to each protein, and ranked the proteins using both the quantitative value and significance of maximal change.

Bar and dot plots of GSEA results were plotted using *ggplot2* package, and heatmaps of GSEA results were made using the *pheatmap* package. Connectivity network (CNET) plots were created using *enrichplot* package where the enriched pathways are represented by their respective colors, and corresponding genes’ adjusted p value. To improve understanding in some of the differential pathways, we performed sub clustering of the pathways. Genes associated with the pathway were extracted from the GO pathway results and subjected to further GO Enrichment Analysis using Reactome/GO molecular function. To refine the results, pathways with a reference gene count exceeding 500 were excluded to avoid overly broad terms, and the remaining pathways were ranked by FDR significance. Starting from the top-ranked pathways, we sequentially included pathways in the CNET plot, excluding those where more than half of the genes overlapped with higher-ranked pathways.

## Ethics approval

Ethical approval was granted by the Sydney Children’s Hospitals Network Human Research Ethics Committee (HREC/18/SCHN/227, 2021/ETH00356). Informed written consent was obtained from all participants.

## Results

### Clinical data

A total of 15 children with a clear history of neurodevelopmental regression (median age 9 (4-14) years at time of sampling, 60% males, with an average time from first regression to sampling of 4.7 years (median 5 (0.5-11) years) and a total of 15 controls (median age 12 (7-16) years, 60% males) were recruited for this study (refer to Table S1). Family history of neurodevelopmental, neuropsychiatric or autoimmune disorders are summarized in Table S2. Parental history of neurodevelopmental or neuropsychiatric disorders was present in 12 of 15 patients (maternal n=10, paternal n=4, including both parental n=2), and an autoimmune condition was present in 8 (maternal n=8, paternal n=2, including both parental n=2). Pregnancy related complications (n=3), infections (n=3) and significant stressors during pregnancy (n=3) were also present (Table S2).

For the 15 patients in this study, 7 had some developmental issues prior to the onset of regression, this included ADHD (n=4), ASD (n=2), and language delay (n=1). The first regression occurred at a median age of 4 (1.5 - 12) years, and 10 children were reported to have triggers associated with the regression event, including infections (n=9, impetigo, Staphylococcal scalded skin syndrome, sore throat, and urinary tract infection), and vaccination (n=1). During the autistic and cognitive regression, symptoms included new onset emotional lability/depression (including anxiety, rage, agitation, irritability) (n=15), new onset or exacerbation of ritualistic and compulsive behaviour (n=15), academic and/or cognitive decline (n=13), loss of language or meaningful words (n=12), behavioural developmental regression or loss in self-help skills (n=12), loss of social skills (n=11), sensory or motor abnormalities (n=11), tics (n=10), loss of imaginative play (n=9), sleep disturbance (n=8) and restricted food intake (n=7). The first regression lasted an average of 11.7 weeks (median 7 weeks, range 2-52 weeks), excluding the two individuals whose symptoms persisted after the initial regression. (Table S2).

The patients had severe neuropsychiatric and behavioural symptoms requiring visits to a hospital emergency department (n=8), and long school absence (>3 months) (n=4). The majority (n=13) reported recurrent regressions (occurring 1-6 times a year), typically triggered by infections, and these deteriorations lasted between 2 and 52 weeks, until subsequent restoration or partial restoration of neurodevelopmental skills (Table S2).

### Blood bulk RNA sequencing

Exploratory bulk RNAseq was performed in 8 children with neurodevelopmental regression compared to 8 healthy controls (Figure 1, Table S1). There was no significant difference in blood cell type composition between patient and control samples (Table S1). Unsupervised principal component analysis (Figure 2A) showed clear discrimination between groups. Top 10 upregulated GSEA GO (FDR <0.05) pathways (Figure 2B, in red) involved immune function (immunoglobulin complex, myeloid cell differentiation/activation, leukocyte migration), epigenetic mechanisms (demethylation), and signaling transduction (secretory granule lumen, endocytosis, tyrosine kinase signaling).

**Figure 2.**
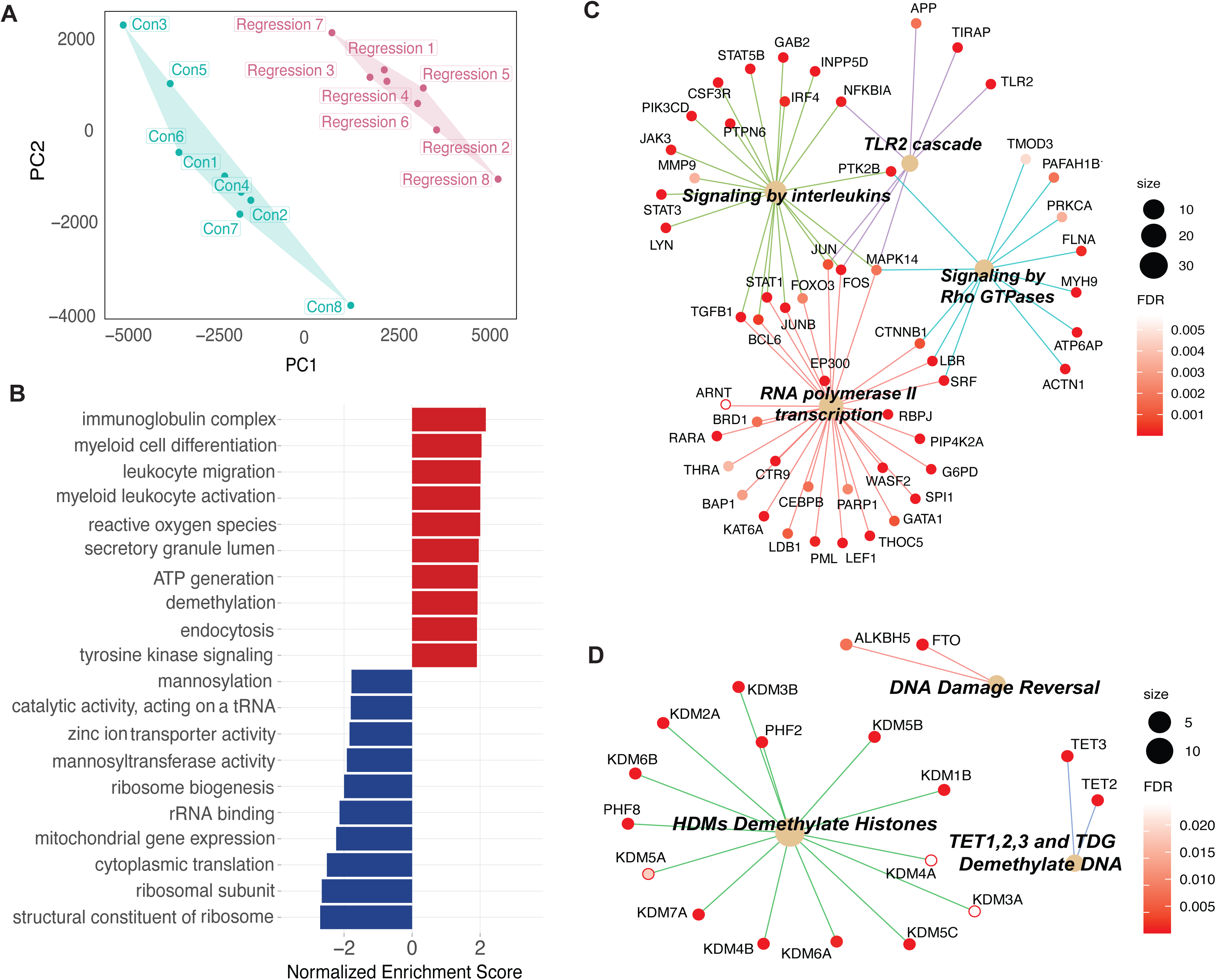
Blood bulk RNA sequencing of children with neurodevelopmental regression versus healthy controls **(A)** Principal component analysis (PCA) performed on peripheral blood bulk RNA sequencing performed in children with neurodevelopmental regression and controls. The x-axis represents Principal Component 1 (PC1), while the y-axis represents Principal Component 2 (PC2). Unbiased hierarchical clustering of gene expression between children with neurodevelopmental regression and controls showed separation of data indicating strong group discrimination post RUV. **(B)** Bar plot of Top 10 up- and down-regulated Gene Set Enrichment Analysis (GSEA) Gene Ontology (GO) bulk RNA sequencing pathways in children with developmental regression compared to the controls. The top 10 up-regulated GSEA GO pathways (red) were mainly involved in immune function (immunoglobulin complex, myeloid cell differentiation/activation, leukocyte migration), epigenetic mechanisms (demethylation), and signaling transduction (secretory granule lumen, endocytosis, tyrosine kinase signaling). The top 10 down-regulated GSEA GO pathways (blue) included ribosomal biogenesis, translation, and mitochondrial expression pathways. **(C)** Genes which enriched the up-regulated myeloid cell differentiation pathway were further clustered via Reactome and included 4 themes including TLR 2 cascade, signaling by interleukins, RNA polymerase II transcription, and signaling by Rho GTPases. **(D)** Genes which enriched the up-regulated demethylation pathway were also further clustered via Reactome and included 3 themes including HDM (histone demethylases), DNA demethylation enzymes (TET enzymes and TDG (thymine DNA glycosylase)), and DNA damage reversal.

Genes which enriched the upregulated myeloid cell differentiation pathway (Figure 2C) were further subclustered via Reactome and included four themes including TLR 2 cascade, signaling by interleukins, RNA polymerase II transcription, and signaling by Rho GTPases. Genes which enriched the upregulated demethylation pathway (Figure 2D) were also further subclustered via Reactome and included three themes including HDMs (histone demethylases including *KDM1B, KDM2A, KDM3A, KDM3B, KDM4A, KDM4B, KDM5A, KDM5B, KDM5C, KDM6A, KDM6B, KDM7A*), DNA demethylation enzymes (TET (ten-eleven translocation), TDG (thymine DNA glycosylase)), and DNA damage reversal.

Top 10 GSEA GO (FDR <0.05) downregulated pathways (Figure 2B, in blue) include ribosomal biogenesis, translation, and mitochondrial expression pathways. Genes which enrich the top 2 downregulated ribosomal pathways include multiple ribosomal genes associated with small subunit (40S) or large subunit (60S) of the eukaryotic and mitochondrial ribosome: *RPS, RPL,* and eukaryotic translation initiation factor genes: *EIF (*Figure S1).

### Blood single-cell RNA sequencing, cohort 1 and cohort 2

We performed two independent cohorts of scRNAseq (Figure 1, each 4 children with neurodevelopmental regression versus 4 controls). These 8 patients were screened for rare pathogenic gene variants that cause ASD and were negative. Cells were clustered into 9 different cell types based on cell markers (Figure 3A), including neutrophils, classical monocytes, natural killer (NK) cells, dendritic cells, basophils, eosinophils, B cells, CD8 T cells, and CD4 T cells.

**Figure 3.**
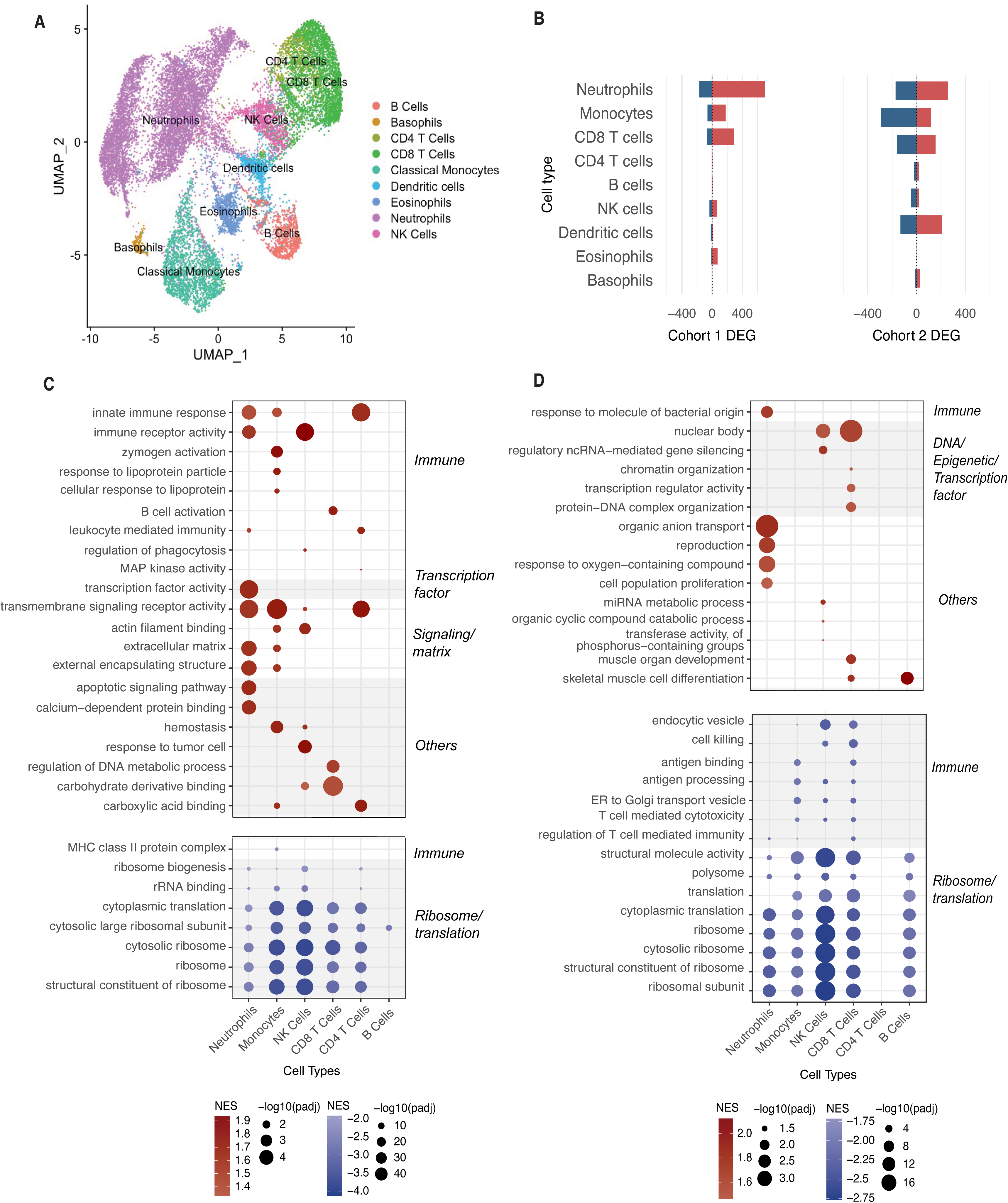
Single-cell blood RNA sequencing (scRNAseq) cohort 1 and 2 of children with neurodevelopmental regression versus controls **(A)** Uniform manifold approximation and projection (UMAP) projection of cohort 1 including 4 children with neurodevelopmental regression and 4 control samples in single-cell transcriptomics identified 9 unique cell types based on cell markers, including neutrophils, classical monocytes, natural killer (NK) cells, dendritic cells, basophils, eosinophils, B cells, CD8 T cells and CD4 T cells. **(B)** Bar chart of differentially expressed genes (DEGs) in individual cell types in cohort 1 (left) and cohort 2 (right), showing genes that are up-regulated (red) or down-regulated (blue) in neurodevelopmental regressive patients compared to controls. **(C)** Dot plot visualizing the top 5 GSEA pathways across all cell types within cohort 1. The dot colour represents the normalized enrichment score (NES), with red indicating upregulation and blue indicating downregulation in neurodevelopmental regressive patients compared to controls, while white represents zero. The size of each dot corresponds to the -log10(padj) of the pathway. Upregulated GSEA GO pathways (FDR <0.05) were clustered in functions involving innate immune (immune response, immune receptor activity), transcription factor, signaling/matrix pathway (transmembrane signaling receptor activity, and actin filament binding). The upregulated innate immune pathways occurred predominantly in neutrophils, monocytes, NK cells and CD4 T cells. The upregulated transcription factor pathways occurred in neutrophils. Downregulated GSEA GO pathways (FDR <0.05) included immune pathways (MHC class II protein complex) and ribosome/translational pathways. Downregulated ribosome/translation pathways were present across most cell types. **(D)** Dot plot visualizing the top 5 GSEA pathways across all cell types within cohort 2. In neurodevelopmental regression patients compared to controls, upregulated GSEA GO pathways (FDR <0.05) had immune functions (response to molecule of bacterial origin), and epigenetic pathways (nuclear body, ncRNA gene silencing activity, chromatin organization, transcription regulator activity, protein-DNA complex organization). Downregulated GSEA GO pathways (FDR <0.05) included immune (endocytic vesicle, cell killing, antigen binding/processing, T cell mediated cytotoxicity/immunity), and ribosome/translational pathways. Downregulated ribosome/translation pathways were present across most cell types (except CD4 T cells).

scRNAseq cohort 1 was performed in 4 children (mean age 9.5 (7-14) years, all males) with neurodevelopmental regression versus 4 healthy controls (11.3 (7-14) years, all males). A total of 7,240 cells from patients and 6,761 cells from controls were sequenced. Differentially expressed genes (DEGs) per cell type ranged from 0 to 868 (Figure 3B, left).

Upregulated GSEA GO pathways (FDR <0.05) (Figure 3C, in red) clustered in functions involving innate immune (immune response, immune receptor activity), transcription factor, and signaling/matrix pathways (transmembrane signaling receptor activity, and actin filament binding). The upregulated innate immune pathways occurred predominantly in neutrophils, monocytes, NK cells and CD4 T cells. The upregulated transcription factor pathways occurred in neutrophils.

Downregulated GSEA GO pathways (FDR <0.05) (Figure 3C, in blue) included immune pathways (MHC class II protein complex) and ribosome/translational pathways. Downregulated ribosome/translation pathways were present across most cell types.

scRNAseq of cohort 2 was performed in a second cohort of 4 children with neurodevelopmental regression (mean age 8.3 (7-10) years, 50% males) versus a second cohort of 4 healthy controls (mean age 10.5 (8-15) years, 50% males). A total of 10,523 cells from patients and 17,237 cells from controls were sequenced. Differentially expressed genes per cell type ranged from 0 to 429 (Figure 3B, right).

Upregulated GSEA GO pathways (FDR <0.05) (Figure 3D, in red) related to immune functions (response to molecule of bacterial origin), and epigenetic pathways (nuclear body, ncRNA gene silencing activity, chromatin organization, transcription regulator activity, protein-DNA complex organization). Downregulated GSEA GO pathways (FDR <0.05) (Figure 3D, in blue) included immune (endocytic vesicle, cell killing, antigen binding/processing, T cell mediated cytotoxicity/immunity), and ribosome/translational pathways. Downregulated ribosome/translation pathways were present across most cell types (except CD4 T cells).

#### PBMC proteomics in cohort 2

The 4 children with neurodevelopmental regression, versus 4 healthy controls who were in the second cohort of scRNAseq, also had proteomics and phosphoproteomics performed (Figure 1). Unsupervised principal component analysis of blood proteomics (Figure S4) showed clear discrimination between groups. A total of 432 upregulated and 720 downregulated differentially abundant proteins were found. The top 5 upregulated GSEA GO pathways (FDR <0.05) (Figure 4A) include receptor-mediated endocytosis and histone acetyltransferase complex. Key proteins that enrich the upregulated receptor-mediated endocytosis pathway (Figure 4B) have functions in signal transduction (*RAB21, EGF),* cytoskeletal dynamics (*ANXA2, TBC1D5),* synaptic function (*FMR1, DTNBP1, HIP1),* cellular stress response *(CLU, SERPINE1, PPT1),* chromatin function *(H1-1)* and mRNA processing (*HNRNPK).* Key proteins that enrich the upregulated histone acetyltransferase complex pathways are involved in chromatin remodelling and transcription regulation (*EP400, ACTL6A, RUVBL1, YEATS*) (Figure S4).

**Figure 4.**
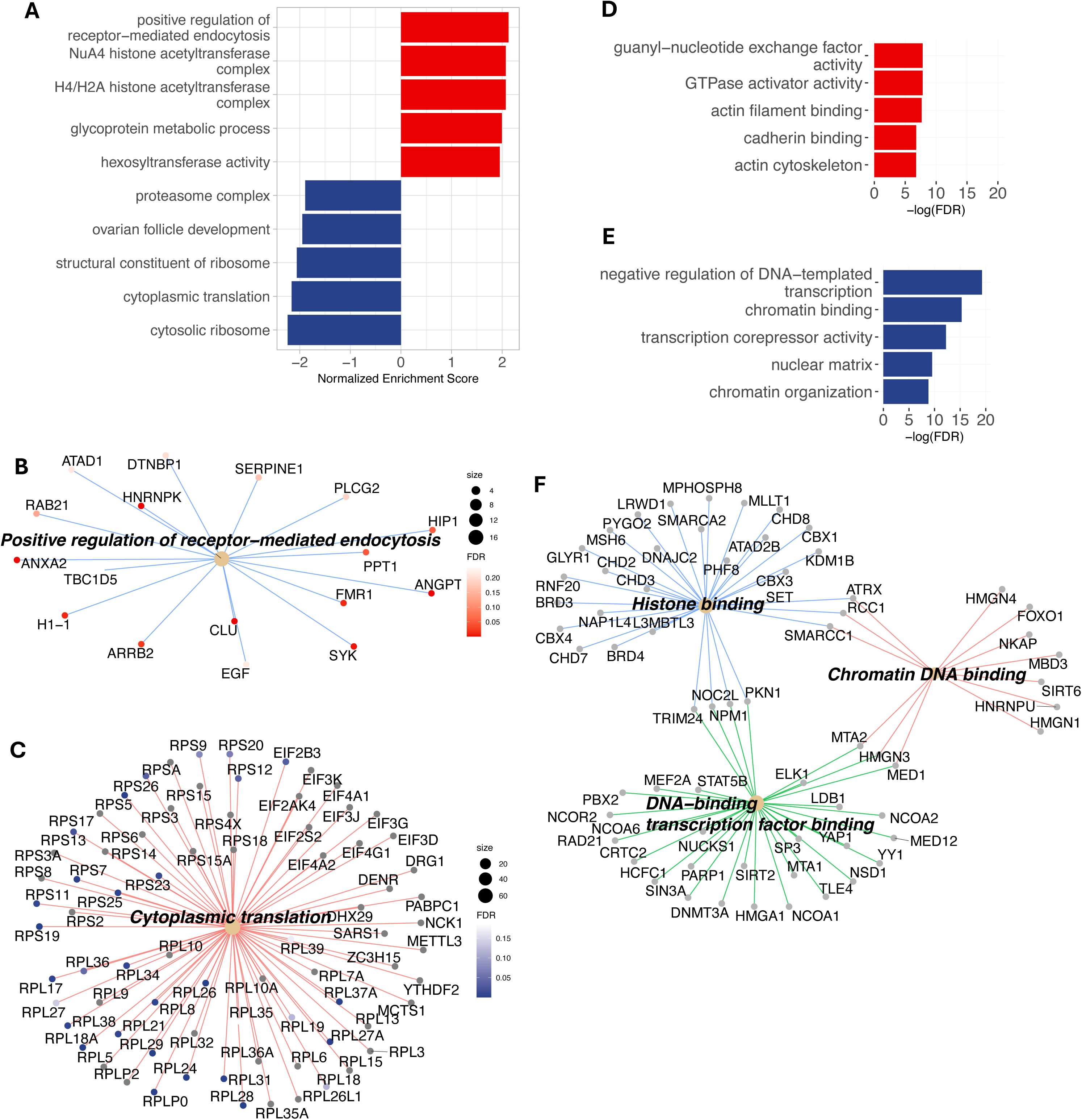
Blood proteomics and phosphoproteomics in cohort 2 of children with neurodevelopmental regression versus healthy controls **(A)** Bar plot of the top 5 up- and down-regulated Gene Set Enrichment Analysis (GSEA) Gene Ontology (GO) proteome pathways in children with developmental regression compared to the controls. The top 2 up-regulated pathways (red) include regulation of receptor-mediated endocytosis, and histone acetyltransferase complex. The top 5 down-regulated pathways (blue) were predominantly ribosomal and translational pathways. **(B)** Connectivity network (CNET) plot of the top up-regulated GSEA GO proteome pathway in patients: positive regulation of receptor-mediated endocytosis. The enriched pathways are represented by their respective colors, and corresponding genes’ adjusted p value. Key proteins that enrich the pathway have functions in signal transduction (*RAB21, EGF),* cytoskeletal dynamics (*ANXA2, TBC1D5),* synaptic function (*FMR1, DTNBP1, HIP1),* cellular stress response *(CLU, SERPINE1, PPT1),* chromatin function *(H1-1), and* mRNA processing (*HNRNPK)*. **(C)** CNET plot of down-regulated GSEA GO proteome pathway in patients: cytoplasmic translation. Key proteins that enrich the pathway include ribosomal genes associated with small subunit (40S) or large subunit (60S) of the eukaryotic ribosome (*RPS, RPL),* eukaryotic translation initiation factor genes (*EIF),* RNA methyltransferase *(METTL)*. **(D)** Bar plot of the top 5 up-regulated over representation analysis (ORA) GO phosphoproteomic pathways in patients compared to controls. The top 5 upregulated ORA GO pathways include guanyl-nucleotide exchange factor activity, GTPase activator activity, actin filament binding, and cadherin binding. **(E)** Bar plot of the top 5 downregulated ORA GO phosphoproteomic pathways in patients compared to controls, include regulation of DNA-templated transcription, transcription corepressor activity, and chromatin binding/organization. **(F)** Key phosphoproteins that enrich the downregulated phosphoproteomic ‘chromatin binding’ pathway was further clustered by GO molecular function pathways and included histone binding (*CHDs, BRDs, KDM1B*), DNA-binding transcription factor binding (*DNMT3A, MEF2A*) and chromatin DNA binding (*HMGNs, HNRNPU*).

Downregulated GSEA GO pathways (FDR <0.05) (Figure 4A) include cytosolic ribosome and translation pathways. Key proteins that enrich the downregulated cytoplasmic translation pathway (Figure 4C) include ribosomal genes associated with small subunit (40S) or large subunit (60S) of the eukaryotic ribosome (*RPS, RPL),* eukaryotic translation initiation factor genes (*EIF),* and RNA methyltransferase *(METTL)*.

### PBMC phosphoproteomics in cohort 2

Unsupervised principal component analysis of blood phosphoproteomics (Figure S5) showed clear discrimination between groups. A total of 2,109 upregulated and 2,396 downregulated differentially abundant phosphopeptides were identified. The top 5 upregulated ORA GO pathways (FDR <0.05) (Figure 4D) include guanyl-nucleotide exchange factor activity, GTPase activator activity, actin filament binding and cadherin binding. Top 5 downregulated ORA GO pathways (FDR <0.05) (Figure 4E) include regulation of DNA-templated transcription, transcription corepressor activity, and chromatin binding/organization. Key phosphoproteins that enrich the downregulated ‘chromatin binding’ pathway (Figure 4F, Figure S5) were further subclustered into GO molecular function pathways including histone binding (*CHDs, BRDs, KDM1B*), DNA-binding transcription factor binding (*DNMT3A, MEF2A*), and chromatin DNA binding (*HMGNs, HNRNPU*).

## Discussion

We conducted in-depth peripheral blood multi-omics profiling in a group of children with neurodevelopmental regression, conditions that significantly impair school functioning and quality of life. Most of the children experienced regression compatible with "autistic regression", although some did not exhibit symptoms that fully met the criteria for an ASD diagnosis. Despite the heterogenous clinical presentations, we identified convergent pathophysiological processes underlying these cases. Through peripheral blood bulk RNA sequencing, single-cell RNA sequencing, and bulk proteomics and phosphoproteomics, we identified key findings. These include dysregulated pathways related to epigenetics (chromatin remodelling, histone modifications, transcription factors) and immune responses, as well as downregulated ribosomal and translational pathways in patients with neurodevelopmental regression. Epigenetic and immune processes are increasingly recognized to play critical roles in the pathogenesis of ASD. Highly penetrant chromatin, histone-related and immune genes are well known contributors to ASD^5–7^. However, genetic testing in our scRNA and proteomic cohorts, including trio whole exome sequencing and autism panel testing, yielded negative results for pathogenic variants associated with autism. However, it is possible that highly penetrant variants in coding or non-coding regions were not captured using this methodology.

Additionally, patients in our study may harbor common vulnerability genes that are missed by clinical genomic screening designed to detect rare pathogenic DNA variations. Thus, the abnormal epigenetic and immune landscape found in our study could arise from susceptibility genes, environmental factors, or an interplay of both factors.

Firstly, we identified a prominent signal of epigenetic dysregulation, involving chromatin organization, histone modifications, and transcription factors. Epigenetic modifications can disrupt critical brain developmental processes, such as neuronal differentiation, migration, synaptic function, and brain connectivity^30^. Modifications such as histone acetylation, methylation, phosphorylation, and ubiquitination play a pivotal role in shaping chromatin structure, directly impacting DNA accessibility to transcriptional machinery. These modifications influence the tightness of nucleosome packaging within chromatin; for instance, increased histone acetylation results in a more relaxed chromatin configuration, which enhances DNA accessibility and promotes gene transcription. In contrast, histone methylation can have dual effects, either loosening or tightening chromatin, depending on the specific methylation pattern, thereby either activating or repressing gene expression. Potential mechanisms that regulate chromatin remodelling include GTPase and actin binding^31^. GTPase is proposed to regulate actin filaments within chromatin remodellers, influencing their interaction with transcription factors, and playing roles in modulating gene regulation^31^. Our phosphoproteomic analysis identified GTPase and actin pathways, however their specific relevance to chromatin modulation in neurodevelopmental regression require further examination. Collectively, these findings point towards the role of epigenetic modifications in neurodevelopment, through pathways which contribute to chromatin remodelling and gene regulation, driving disruptions in brain development in these children.

Secondly, dysregulated ribosomal and translational pathways were consistently identified through the multi-omics profiling in our cohort of children with neurodevelopmental regression. The translation of mRNA into functional proteins occurs in ribosomes, which play critical roles in brain development. Disruptions in the translational machinery have been linked to NDDs^32,33^. Monogenic mutations affecting components of the translational process, such as ribosomal proteins (ribosomopathies), are associated with NDDs^32^. Additionally, environmental exposures, such as infections and stressors, can impair ribosomal biogenesis, through epigenetic processes^34–36^. During stress conditions, such as proteostasis imbalance, nutrient deprivation, and oxidative stress, cells have been shown to assemble heterochromatin in rDNA, leading to its transcriptional repression^37,38^.

Perturbations in ribosomal biogenesis can reduce protein synthesis crucial for dendritic and synaptic function, contributing to changes in neuronal connectivity associated with NDDs. In our study, the consistent downregulation of ribosomal and translational pathways observed across bulk RNA sequencing, single-cell RNA sequencing, and proteomic analyses highlights the importance of these pathways. These findings align with translational-immune signatures reported in peripheral blood and brain transcriptomic studies of more classical NDDs^39^. Leukocyte RNA expression from two independent cohorts of ASD boys aged 1 to 4 years (142 discovery participants and 73 replication participants) using Illumina microarrays, showed differentially co-expressed genes enriched in translation and immune/inflammation functions^39^. The gene signature observed in our cohort with neurodevelopmental regression suggests shared underlying pathophysiological mechanisms between regressive and non-regressive NDD phenotypes. Together, we propose that ribosomal protein dysregulation is a central feature in NDDs and a potential marker of abnormal gene (epigenetic) regulation^18^.

Thirdly, we identified dysregulated immune pathways in RNA and proteome sequencing in our cohorts. Single-cell RNA sequencing revealed heterogenous patterns of immune regulation across cell types, with a mix of up- and downregulation instead of a uniform trend. Immune dysregulation has also been observed in various NDDs, through studies of peripheral blood and brain samples in humans^40–42^. In peripheral blood clinical studies, abnormal cytokine profiles, immunoglobulin levels, immune cell subpopulations, and gene expression profiling of lymphocytes have been elucidated in ASD, ADHD and Tourette syndrome^40,42^.

Several molecular signaling pathways, including those involving cytokines, major histocompatibility complex molecules, complement factors, and microglia, have been identified as links between peripheral and central immune activation, and the development of NDD phenotypes^40^. Several ASD risk genes are involved in immune functions and the transcriptional regulation of immune networks^43^. Analysis of transcription factor networks in differentially expressed genes from brain tissue and peripheral mononuclear cells in ASD has identified *RELA* and *IRF* as key targets, both of which play crucial roles in regulating the immune system^44^.

Epigenetic modifications and immune dysregulation in NDDs can also arise from environmental influences. Environmental influences on child brain development begins in the pre-natal period and continues postnatally. Animal models and human epidemiological data support an association between maternal inflammation during pregnancy and offspring NDDs^2,45^. In our study, there was high prevalence of maternal autoimmunity, psychiatric disorders (e.g. anxiety, depression) and pregnancy related complications (e.g. gestational diabetes, infections). These diverse maternal conditions during pregnancy lead to inflammatory perturbations to the *in utero* environment, and have been shown to adversely affect fetal neurodevelopment and increase the risk of NDDs in human offspring^2,46^. A meta-analysis of two high-risk prospective human cohort studies involving umbilical cord blood revealed transcriptional changes to chromatin, autoimmune, and environmental response genes in children with NDDs^47^. This suggests that epigenetic factors and immune processes play important roles at the gene-environment interface early in the development of NDDs. Chromatin genes have high expression during gestation and are downregulated after birth. This observation supports the window of vulnerability hypothesis, in which the early stages of brain development are the most sensitive to perturbations, and disruptions during the critical periods can affect neurodevelopment. Additionally, animal models show that immune insults during pregnancy can prime microglia via epigenetic reprogramming, heightening susceptibility to postnatal “second hit” events in offspring, such as infections or stress, which can trigger neurobehavioral abnormalities^48,49^. In our cohort, infection provoked regressions were highly prevalent, often recurrent, and had significant lasting effects on child development. While some children were able to recover between regressions, others experienced progressive symptoms without return to developmental baseline. Postnatal immune challenges can induce peripheral immune activation, which communicates with brain immune cells through central-peripheral cross-talk, triggering microglia to adopt an activated state and heightened pro-inflammatory response^49,50^. Post-mortem ASD brain studies reveal upregulation of immune gene expression in microglial markers, corresponding to hypomethylation of these genes, postulated to be driven by environmental factors^51,52^. Collectively, studies indicate that epigenetic and immune pathways represent a key convergence point for the diverse genetic and environmental risk factors associated with NDDs throughout life.

The strengths of our study include an in-depth multi-omics analysis in children with autistic and cognitive regression, revealing consistent epigenetic, transcriptional, immune, and ribosomal abnormalities. We hypothesize that epigenetic modifications in brain and blood immune cells contribute to downstream transcriptional, immune, and ribosomal dysregulation, all of which are critical for normal brain development. Limitations of this study include small sample size and heterogeneity in clinical symptomatology. Clinical histories of regression were sometimes obtained retrospectively, which may introduce recall bias. However, extensive efforts were made to corroborate these accounts through video evidence and observations from multiple independent sources (e.g. teachers). Secondly, there is absence of standardised definitions of neurodevelopmental regression, despite ongoing efforts to standardize and validate these criterias^15^. Thirdly, omics analyses were performed many years after initial regression event, and the majority of the children were taking psychiatric medications (including anti-depressants) during this study, which can theoretically influence gene and protein expression. Fourthly, this study focused on children with loss of skills or developmental regression, and did not include more classical presentations of NDDs for comparison. As a result, direct comparisons of gene expression profile between our cohort and more classical ASD, OCD or tics/Tourette syndrome was not possible. Fifthly, it is important to highlight that bulk and HIVE single-cell RNA sequencing of whole blood included all leukocyte populations, whereas proteomics and phosphoproteomics were performed on PBMCs (excluding neutrophils). Also, we performed investigations only in blood and not in brain. In clinical cohorts like this, omics analysis of brain tissues is not feasible, making investigations using peripheral blood a more practical and accessible approach. We have shown that multi-omic analysis of peripheral blood has potential clinical utility in distinguishing between patients and controls. Our results compliment previous findings of dysregulated epigenetic, chromatin, and immune pathways in brain tissue of individuals with neurodevelopmental disorders. We propose that peripheral blood RNA and proteomic sequencing can effectively capture the interactions between genetic and environmental factors that are critical for the expression of NDDs, offering valuable insights into their underlying pathogenesis.

Future studies involving larger, treatment-naïve cohorts of NDDs, together with careful evaluation of genetic backgrounds and environmental exposures, will enable further genotype-phenotype correlations, including those across different developmental trajectories. Previous investigators have proposed that the relative genetic and environmental contributions to ASD include rare pathogenic de novo variants (9.5%), rare pathogenic inherited variants (2.6%), common inherited variants (49.8%) and environmental factors (38.1%)^14^. Exploring common gene variants and utilizing polygenic scores could provide valuable insights into whether our omics signature is associated with common genetic variants, environmental factors, or a combination of both. This approach will help identify convergent or divergent pathways associated with these different developmental phenotypes, which will help to develop targeted therapeutics for these different subgroups of patients. Omics analyses should also be conducted at different time points in the disease, to examine the stability of peripheral blood genes and protein expression. Additionally, future studies should incorporate concurrent epigenetic profiling, including ATAC sequencing, histone modifications, and methylation to further elucidate underlying epigenetic modifications for these gene expression perturbations. While our study highlights epigenetic mechanisms, the timing and triggers of epigenetic modifications in these children remain unclear. These changes could stem from heritable epigenetic marks from their parents or may have occurred during pregnancy or early life. To date, treatments for NDDs primarily focus on managing symptoms and providing developmental support. However, therapies that target immune dysregulation or epigenetic modifications, such as butyrate, may offer effective treatment options for these patients^53,54^.

## Resource availability

Anonymized data not published within this article and R code will be made available by request from any qualified investigator.

## Data Availability

All data produced in the present study are available upon reasonable request to the authors.

## Acknowledgements

We would like to thank the patients and their families for participating in this study. We would like to thank the doctors and nurses in the Department of Paediatric Endocrinology at The Children’s Hospital at Westmead, Sydney for their help and support to recruit controls. We would like to thank Children’s Medical Research Institute for access to the Biomedical Proteomics Facility and bioinformatic support and thank the Australian Genome Research Facility for providing expertise in sequencing.

## Author contributions

Conceptualization: HN, VXH, SP, RCD; Methodology: HN, VH, SP, RCD; Investigation: HN, VXH, BK, KZ, SA, SP; Formal analysis: HN, VXH, KZ, BG, NA, LLM, XL, RD, MG, SP, RCD, Writing – Original Draft: HN, VH, RCD; Writing – Review & Editing: MG, SP, RCD; Funding Acquisition, RCD; Supervision: SP and RCD.

## Supplemental information

Table S1: Children with neurodevelopmental regression and healthy controls

Table S2: Clinical characteristics of children with neurodevelopmental regression

Table S3: Bulk RNA sequencing pathway analysis

Table S4: Single-cell RNA seq pathway analysis – Cohort 1

Table S5: Single-cell RNA seq pathway analysis – Cohort 2

Table S6: Proteomics pathway analysis

Table S7: Phosphoproteomics pathway analysis

Figure S1: Exploratory bulk RNA seq

Figure S2: Single-cell RNA seq analysis – Cohort 1

Figure S3: Single-cell RNA seq analysis – Cohort 2

Figure S4: Proteomics analysis – Cohort 2

Figure S5: Phosphoproteomics analysis – Cohort 2

