## Supplemental Figure 1-5 for "Chromatin, transcriptional and immune dysregulation in children with neurodevelopmental regression"

### Slide 1
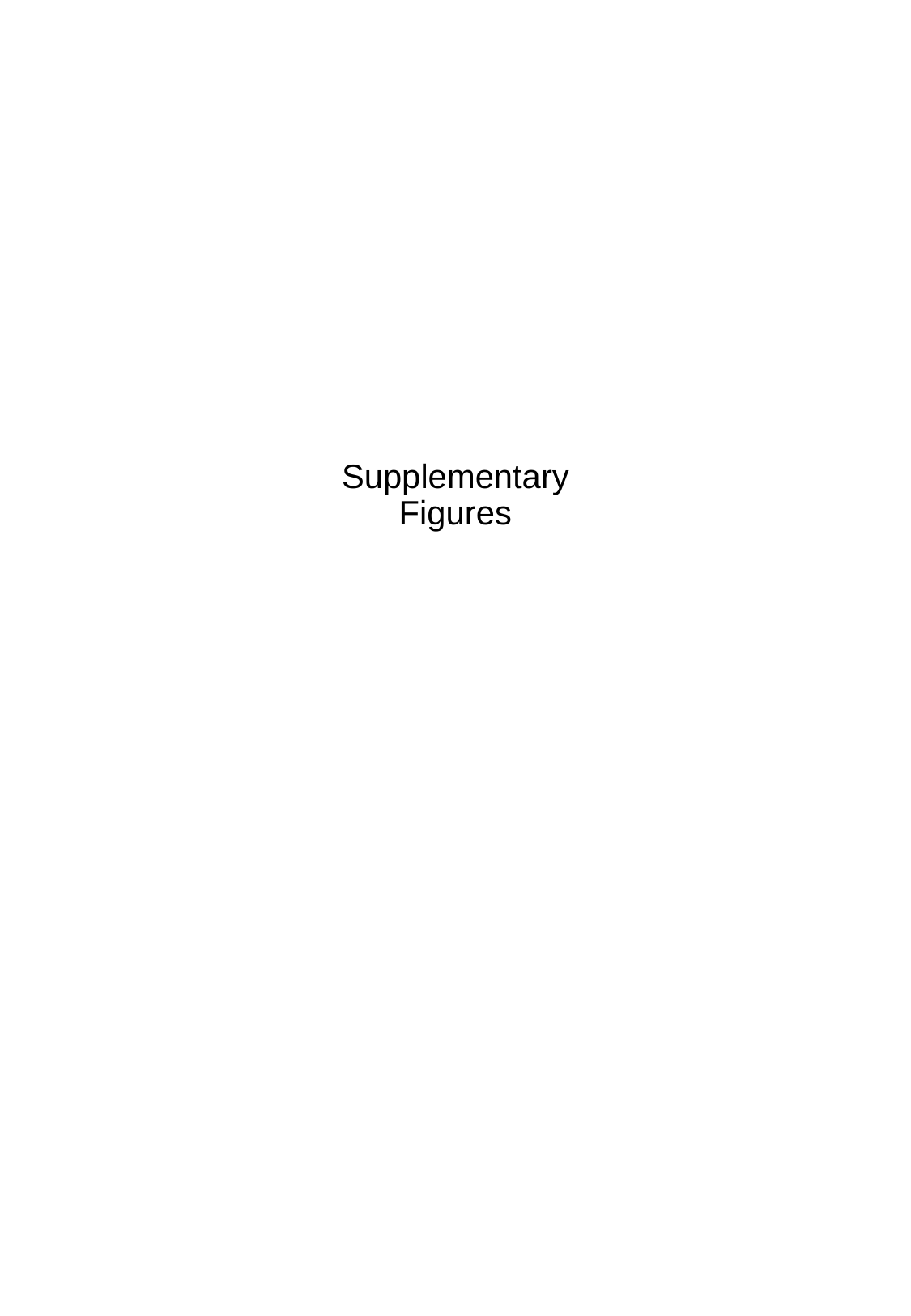

#### Supplementary Figures

### Slide 2
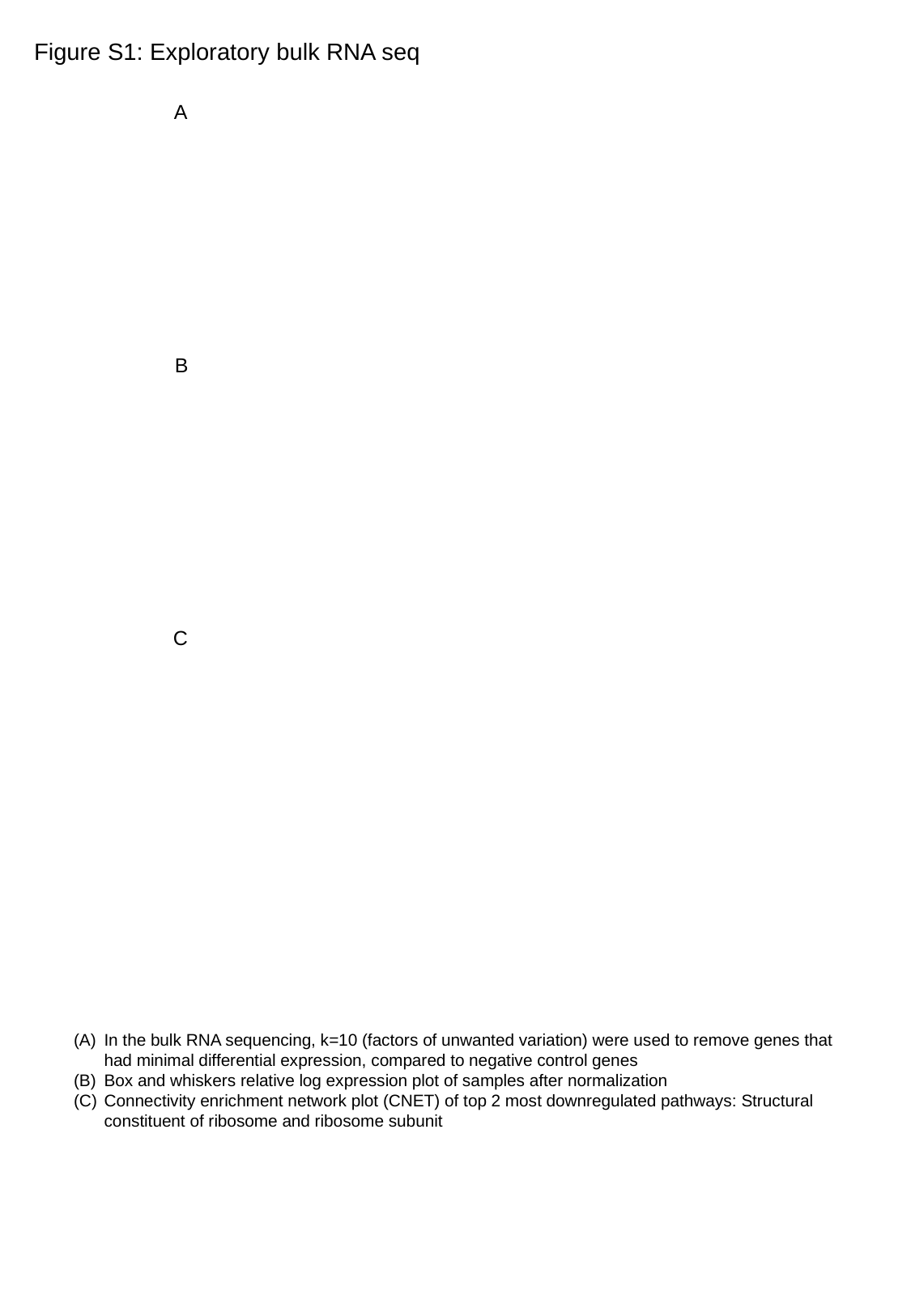

Figure S1: Exploratory bulk RNA seq
A
B
C
In the bulk RNA sequencing, k=10 (factors of unwanted variation) were used to remove genes that had minimal differential expression, compared to negative control genes
Box and whiskers relative log expression plot of samples after normalization
Connectivity enrichment network plot (CNET) of top 2 most downregulated pathways: Structural constituent of ribosome and ribosome subunit

### Slide 3
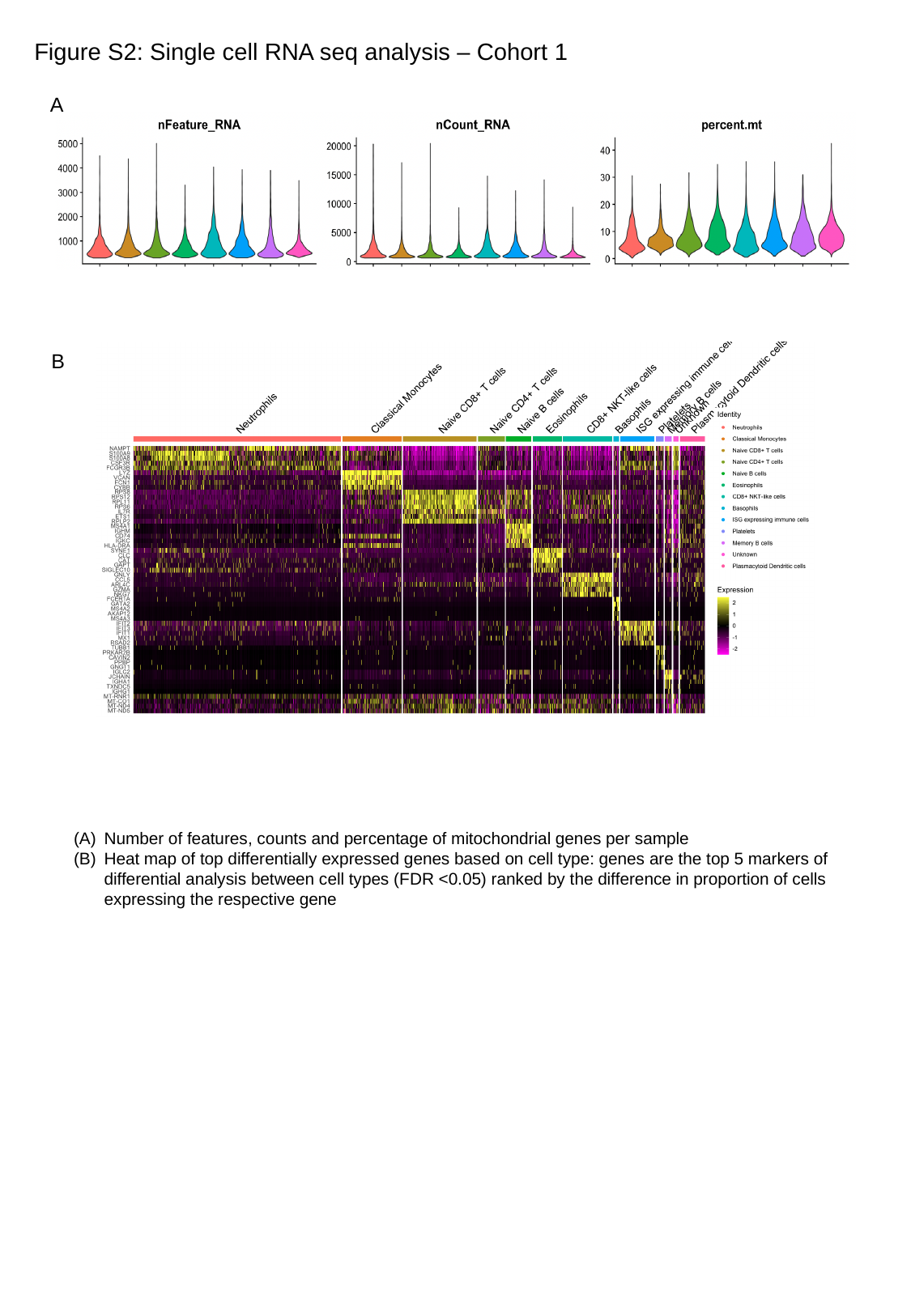

Figure S2: Single cell RNA seq analysis – Cohort 1
A
B
Number of features, counts and percentage of mitochondrial genes per sample
Heat map of top differentially expressed genes based on cell type: genes are the top 5 markers of differential analysis between cell types (FDR <0.05) ranked by the difference in proportion of cells expressing the respective gene

### Slide 4
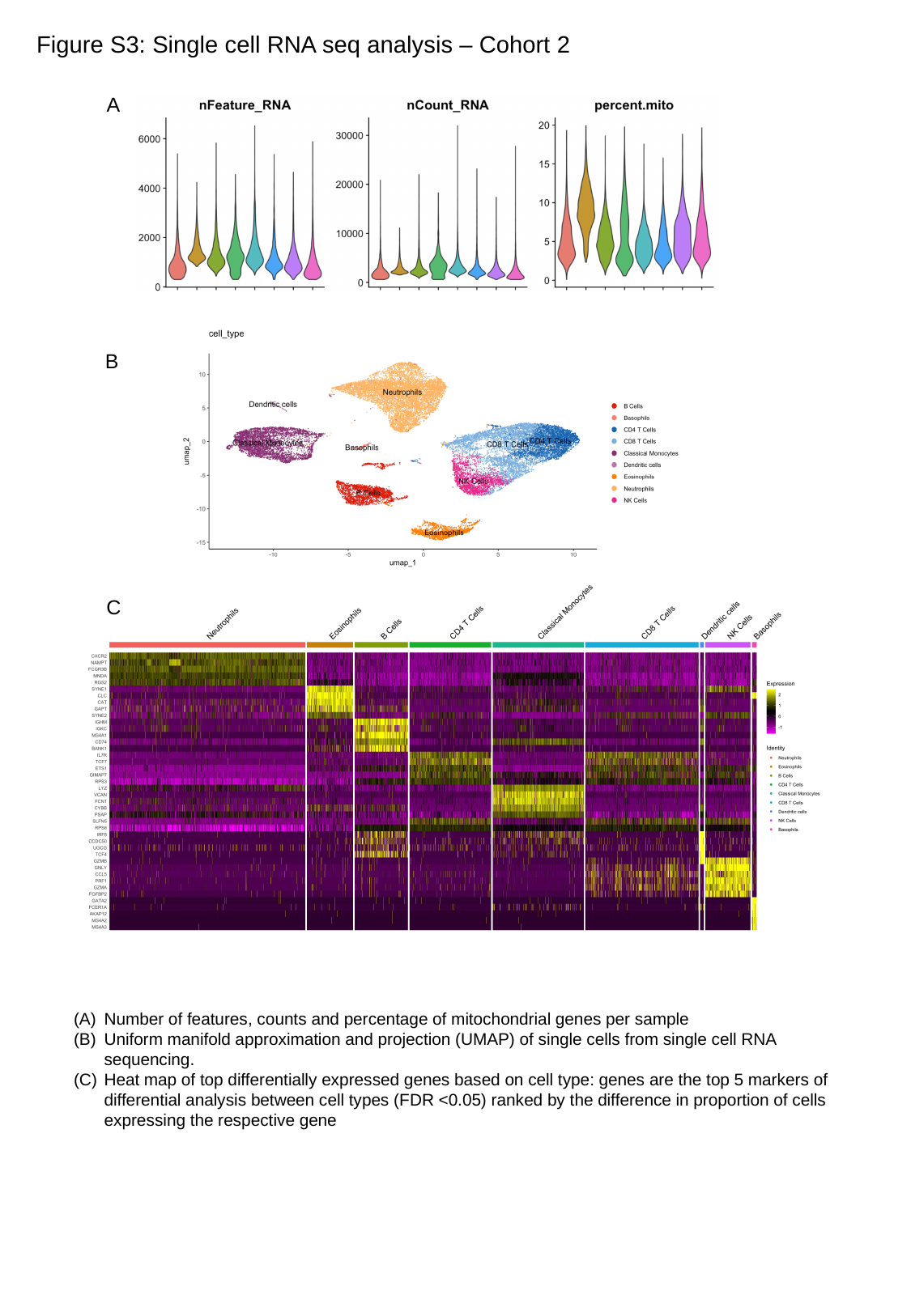

Figure S3: Single cell RNA seq analysis – Cohort 2
A
B
C
Number of features, counts and percentage of mitochondrial genes per sample
Uniform manifold approximation and projection (UMAP) of single cells from single cell RNA sequencing.
Heat map of top differentially expressed genes based on cell type: genes are the top 5 markers of differential analysis between cell types (FDR <0.05) ranked by the difference in proportion of cells expressing the respective gene

### Slide 5
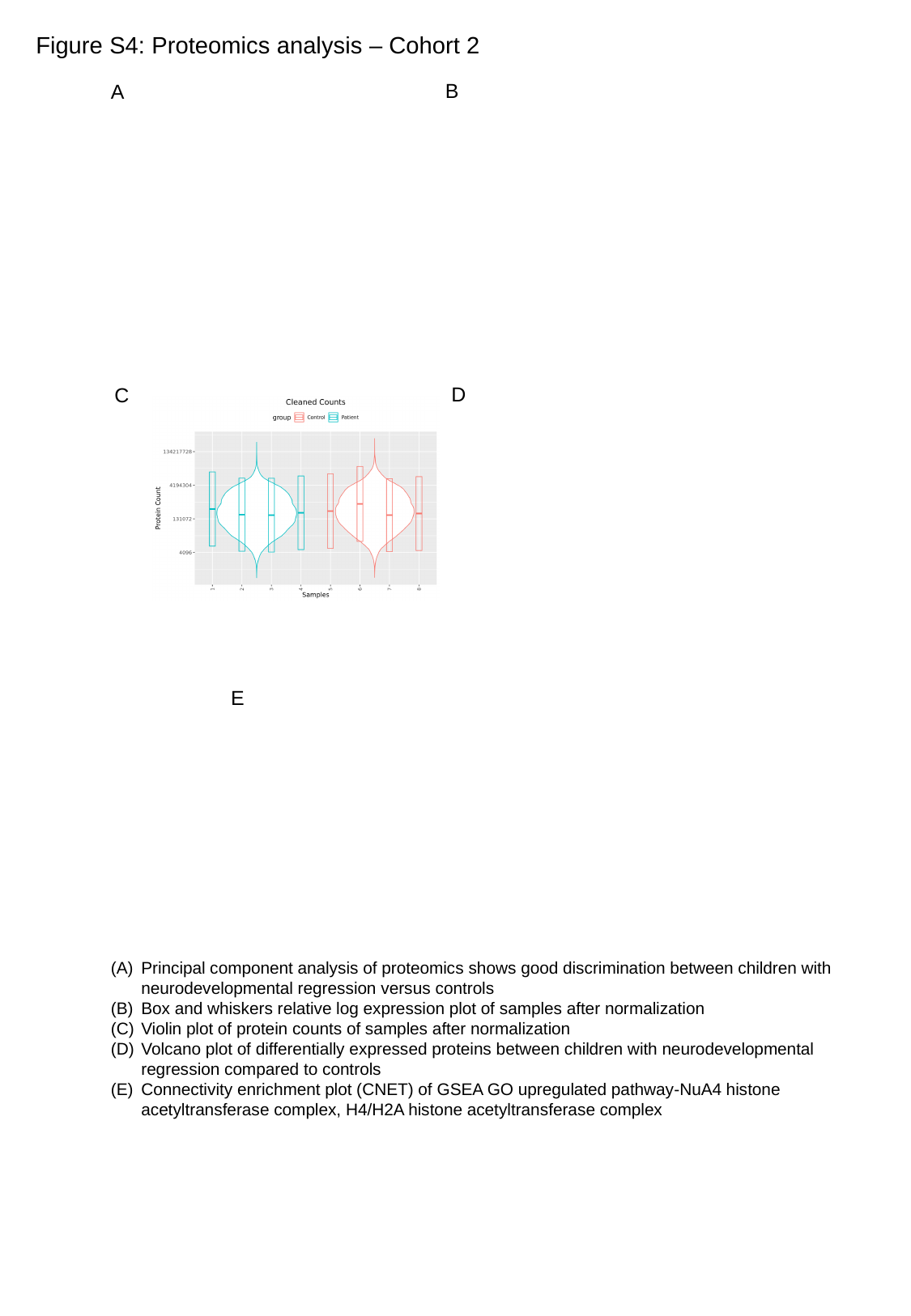

Figure S4: Proteomics analysis – Cohort 2
B
A
D
C
E
Principal component analysis of proteomics shows good discrimination between children with neurodevelopmental regression versus controls
Box and whiskers relative log expression plot of samples after normalization
Violin plot of protein counts of samples after normalization
Volcano plot of differentially expressed proteins between children with neurodevelopmental regression compared to controls
Connectivity enrichment plot (CNET) of GSEA GO upregulated pathway-NuA4 histone acetyltransferase complex, H4/H2A histone acetyltransferase complex

### Slide 6
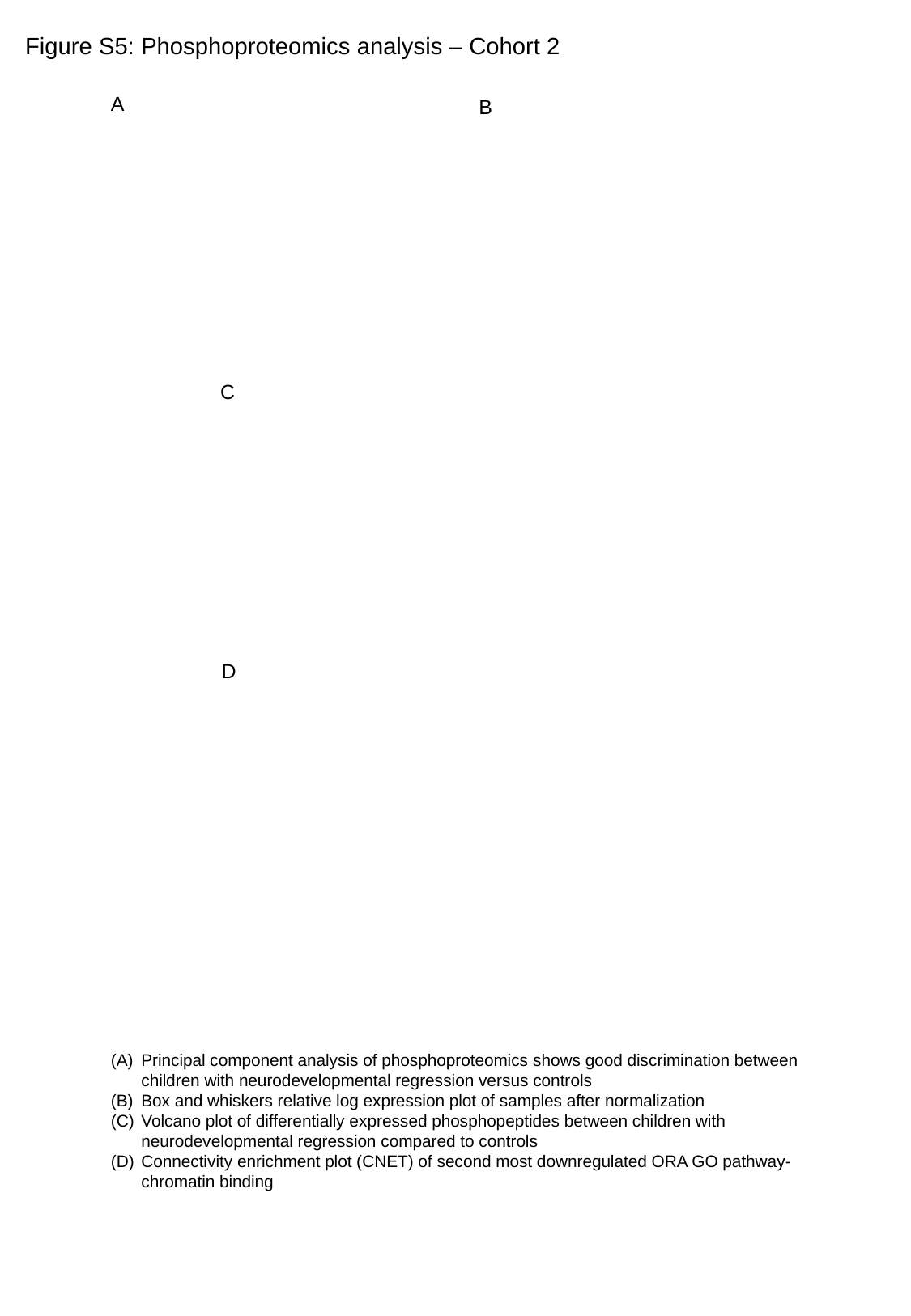

Figure S5: Phosphoproteomics analysis – Cohort 2
A
B
C
D
Principal component analysis of phosphoproteomics shows good discrimination between children with neurodevelopmental regression versus controls
Box and whiskers relative log expression plot of samples after normalization
Volcano plot of differentially expressed phosphopeptides between children with neurodevelopmental regression compared to controls
Connectivity enrichment plot (CNET) of second most downregulated ORA GO pathway- chromatin binding
