## Supplemental Table 1-2 for "Chromatin, transcriptional and immune dysregulation in children with neurodevelopmental regression"

Table S1: Clinical cohort: Demographics, omics analyses, full blood count results, and medications taken at time of omics testing

| **Patients** | **Gender** | **Age** | **Time from first regression to sampling** | **Bulk RNA seq** | **scRNA seq (cohort)** | **Proteome** | **Phospho-proteomics** | **White cell count**  **(x 10^9^/L)** | **Neutrophils**  **(x 10^9^/L)** | **Lymphocytes**  **(x 10^9^/L)** | **Monocytes**  **(x 10^9^/L)** | **Medications taken at the**  **time of blood draw** |
| --- | --- | --- | --- | --- | --- | --- | --- | --- | --- | --- | --- | --- |
| 1 | M | 10-14 | 7y |  | X (1) |  |  | 13.1 | 8 | 3 | 1.1 | Risperidone, Aripiprazole, Fluoxetine, Propranolol |
| 2 | M | 5-9 | 5y |  | X (1) |  |  | 11.4 | 7.9 | 1.9 | 1 | Dexamphetamine, Guanfacine, Quetiapine, Fluoxetine, Melatonin, Cyproheptadine |
| 3 | M | 5-9 | 5y |  | X (1) |  |  | 7.6 | 2.8 | 3.7 | 0.5 | Aripiprazole, Sertraline |
| 4 | M | 5-9 | 5.5y | x | X (1) |  |  | 6.1 | 2.2 | 2.7 | 0.5 | Risperidone, Guanfacine |
| 5 | M | 5-9 | 2y |  | X (2) | X | X | 7 | 2.4 | 4.1 | 0.3 | Guanfacine, Lisdexamfetamine, Clonidine |
| 6 | M | 5-9 | 2y |  | X (2) | X | X | 7 | 3 | 3.1 | 0.6 | Guanfacine |
| 7 | F | 10-14 | 7y |  | X (2) | X | X | 10.9 | 5.8 | 3.9 | 0.8 | Nystatin, Cholestyramine, Osmolax |
| 8 | F | 5-9 | 5y |  | X (2) | X | X | 6.6 | 1.6 | 4.5 | 0.4 | Topiramate, Lamotrigine, Guanfacine, Clonidine, Aripiprazole |
| 9 | M | 10-14 | 9y | x |  |  |  | 5.8 | 2.5 | 2.5 | 0.6 | Erythromycin, Amantadine, Melatonin, Promethazine |
| 10 | M | 5-9 | 3y | x |  |  |  | 5.7 | 3.4 | 1.5 | 0.3 | Lisdexamfetamine, Guanfacine, Fluoxetine |
| 11 | M | 10-14 | 2y | x |  |  |  | 10.6 | 6.3 | 3.2 | 0.5 | Aripiprazole, Diazepam, Clonidine Valproate, Lithium |
| 12 | F | 10-14 | 6y | x |  |  |  | 10.3 | 6.7 | 2.7 | 0.6 | Fluoxetine, Clonidine, Melatonin Risperidonem Propranolol |
| 13 | F | 5-9 | 1y | x |  |  |  | 7.7 | 3.6 | 3.1 | 0.5 | None |
| 14 | F | 0-4 | 0.5y | x |  |  |  | 8.7 | 3.9 | 3.8 | 0.7 | Trimethoprim/sulfamethoxazole |
| 15 | F | 10-14 | 11y | x |  |  |  | 10.6 | 6.6 | 3.2 | 0.5 | None |

| **Controls** | **Gender** | **Age** | **Bulk RNA** | **scRNA**  **seq (Cohort)** | **Proteome** | **Phospho-proteomics** | **White cell count**  **(x 10^9^/L)** | **Neutrophils**  **(x 10^9^/L)** | **Lymphocytes**  **(x 10^9^/L)** | **Monocytes**  **(x 10^9^/L)** |
| --- | --- | --- | --- | --- | --- | --- | --- | --- | --- | --- |
| 1_1 | M | 5-9 |  | X (1) |  |  | 7.4 | 3 | 3.1 | 0.6 |
| 1_2 | M | 5-9 |  | X (2) | X | X | 7.8 | 3.6 | 2.7 | 0.6 |
| 2 | M | 10-14 | X | X (1) |  |  | 7.1 | 2.7 | 3.6 | 0.3 |
| 3 | M | 10-14 |  | X (1) |  |  | 11 | 7.3 | 2.4 | 1.2 |
| 4_1 | M | 10-14 |  | X (1) |  |  | 7 | 2.1 | 3.6 | 0.5 |
| 4_2 | M | 15-19 |  | X (2) | X | X | 6.9 | 2.3 | 3.5 | 0.3 |
| 4_3 | M | 10-14 | X |  |  |  | 6.7 | 2.5 | 2.8 | 0.4 |
| 5 | F | 10-14 |  | X (2) | X | X | 7.5 | 3.1 | 2.7 | 0.7 |
| 6 | F | 5-9 |  | X (2) | X | X | 5.1 | 1.9 | 2.2 | 0.5 |
| 8 | F | 15-19 | X |  |  |  |  |  |  |  |
| 9 | F | 10-14 | X |  |  |  | 8.7 | 4.1 | 3.3 | 0.9 |
| 10 | F | 10-14 | X |  |  |  | 8.9 | 7.1 | 1.1 | 0.7 |
| 11 | F | 10-14 | X |  |  |  | 5.6 | 3.4 | 1.7 | 0.4 |
| 12 | M | 10-14 | X |  |  |  | 7.5 | 3.7 | 3 | 0.6 |
| 13 | M | 15-19 | X |  |  |  | 9.7 | 4.7 | 3.6 | 1 |

Table S2: Children with neurodevelopmental regression: family history, pregnancy history, regression details, clinical phenotype, and diagnoses

|  |  |  |  |  | Clinical phenotype | | | | | | |  |  |  |
| --- | --- | --- | --- | --- | --- | --- | --- | --- | --- | --- | --- | --- | --- | --- |
| Case | Family history | Pregnancy (maternal) history | Age at regression,  Pre-regression development | Trigger of the first regression | Loss of Social Skills | Loss of Language | Academic/cognitive Decline | OCD, Repetition | Anxiety/ED | ADHD Traits | Behavioural regression | Duration of the first regression and regaining of previous skills | Trajectory of symptoms | Current diagnosis  (Duration of follow up) |
| 1 | ADHD, HT (M), Sacroiliitis (F); Depression (B) | - | 5-9y ASD | Sore throat | + | + | + | + | + | + | + | 2 months, partially regained skills | RR | ASD, ADHD, OCD, Tourette, anxiety (9y) |
| 2 | Depression, anxiety, PTSD, HT (M) | - | 0-4y Normal | Pneumonia | + | + | + | + | + | + | + | 1–2 months, never regained skills | RR | ASD, ADHD, OCD, anxiety, eating restriction (7y) |
| 3 | Depression, anxiety, PTSD, Grave's disease, MS (M); Crohn's disease (F) | UTI | 0-4y Normal | Minor head bump | - | + | + | + | + | - | + | 1 month, never regained skills | SF | OCD, depression (7y) |
| 4 | Anxiety, depression, HT, coeliac disease, psoriasis (M) | Severe work-related stress | 0-4y Normal | - | + | + | + | + | + | + | + | Symptoms persisted, never regained skills | RR | ASD, ADHD, mod LD, anxiety, compulsion (9y) |
| 5 | ADHD, anxiety (M); ASD, mild LD, ADHD (B) | Pneumonia | 5-9y ADHD | URI | + | + | + | + | + | + | + | 2–3 months, partially regained skills | SF | ASD, ADHD, OCD, LD, tics, insomnia (4y) |
| 6 | - | Pre-eclampsia, obesity, GDM, HG, delivered at 35 weeks | 5-9y ADHD | Impetigo | + | + | + | + | + | + | - | Few months, never regained skills | CP | ASD, ADHD, low average intelligence, ED (5y) |
| 7 | - | Mild GDM | 0-4y Normal | Sore throat, dry cough | + | + | + | + | + | + | + | >6 months, never regained skills | MF | ASD, ADHD, OCD, insomnia, ED (8y) |
| 8 | Schizophrenia (F) | - | 0-4y Normal | AGE, tonsilitis and URI | + | - | + | + | + | + | + | Few months, never regained skills | SF | ASD, inattention, OCD, anxiety, insomnia, ED (6y) |
| 9 | Anxiety, chronic fatigue syndrome (M) | Preterm labour at 27 weeks, delivered at 34 weeks (BW 2kg), 2-week NICU | 0-4y Language delay | Stress – sibling transported by ambulance | - | + | + | + | + | + | + | 2–6 weeks, partially regained skills | RR | ASD, anxiety, depression, dyspraxia, LD, working memory dysfunction (12y) |
| 10 | Mild ID, ADHD, OCD, coeliac disease (M); Depression, anxiety, PTSD (F, M, B); ADHD (B) | Stress, uncontrolled asthma, recurrent UTI, delivered at 33 weeks; | 5-9y Normal | UTI | + | + | + | + | + | + | + | Symptoms persisted, never regained skills | SF | ASD, ADHD, OCD, anxiety (5y) |
| 11 | OCD (M); anxiety, depression (M, F); ADHD (B); | - | 10-14y ASD | - | + | + | + | + | + | - | + | Years, never regained skills, slow decline | CP | ASD, mild ID, Tourette, OCD, anxiety (5y) |
| 12 | Anxiety, Graves disease (M); ADHD, tics (M, F) | - | 0-4y Hyperactivity | - | + | - | + | + | + | + | - | Symptoms persisted, never regained skills | MF | ASD, ADHD, Tourette, ODD, anxiety, OCD (7y) |
| 13 | Anxiety, postnatal depression (M) | Unexplained microscopic haematuria | 0-4y Hyperactivity | Croup | - | - | - | + | + | + | - | 4 weeks, partially regained skills | RR | ADHD, OCD, anxiety, tics (5y) |
| 14 | - | Stress due to 2 bereavements | 0-4y Normal | SSSS | + | + | + | + | + | + | + | 1 month, never regained skills | MF | ASD, ADHD, OCD, Tourette, anxiety (4y) |
| 15 | Tics (F); Vitiligo (M) | Type 2 diabetes, polycystic ovary syndrome | 0-4y Normal | Influenza vaccine | - | + | - | + | + | + | + | Few weeks, partially regained skills | MF | Tourette, ADHD, OCD, anxiety (14y) |

ADHD: attention-deficit/hyperactivity disorder, AGE: acute gastroenteritis, ASD: autism spectrum disorder, B: brother/brothers, BW: birth weight, CP: chronic progressive, ED: emotional dysregulation, F: father, GDM: gestational diabetes mellitus, HG: hyperemesis gravidarum, HT: Hashimoto thyroiditis, ID: intellectual disability, LD: learning disability, M: mother, MF: moderate fluctuations, MS: multiple sclerosis, NDD: neurodevelopmental disorder, NICU: neonatal intensive care unit, OCD: obsessive-compulsive disorder, PTSD: post-traumatic stress disorder, SF: severe fluctuations, SSSS: staphylococcal scalded skin syndrome, TBI: traumatic brain injury, ODD: oppositional-defiant disorder, RR: relapsing remitting, URI: upper respiratory infection, UTI: urinary tract infection.
